# Minimum Physical Activities Protective Against Alzheimer’s Disease in Late Life: A Systematic Review

**DOI:** 10.1101/2025.04.01.25325006

**Authors:** Amy Sakazaki, Austin Lui, Haneef Muhammad, Claire Vu, Aiden le Roux, Patty Lacayo, Shin Murakami

## Abstract

**Introduction:** Previous studies indicate an inverse relationship between physical activity (PA) and the risk of Alzheimer’s disease (AD). While they highlighted the health benefits of PA, the specific effects of PA in late life remain unclear, and intense PA may be challenging for older adults. Moreover, there is significant variation in how PA is assessed, including the timing and types of activities considered.

**Objective:** This review aims to evaluate existing literature to determine the effects of PA with an emphasis on late life PA and the minimum levels for older adults.

**Methods:** We conducted a systematic review via PRISMA protocol using MEDLINE and CINAHL databases, last assessed in July 2023. Studies that met inclusion criteria were prospective cohort or interventional studies, written in English, and measured physical activity in a cohort who did not have dementia, AD, or cognitive decline at baseline. Retrospective cohort, cross-sectional, case reports, and studies not meeting the inclusion criteria were excluded. Each study was evaluated in 7 domains of bias using the Robins-E tool.

**Results:** Out of 2,322 studies screened, 17 met the inclusion criteria, including six new studies not included in the previous systematic review. This resulted in 206,463 participants from North America (United States and Canada) and Europe (Denmark, Finland, Italy, Sweden, and the United Kingdom). Our method effectively reduced the number of duplicated studies during screenings, resulting in 92 duplications compared to 3,580 in the previous review. The risk of bias assessment in the quality of evidence was low risk in 13 studies and some concerns in four studies. Four studies assessed PA at mid-life (average age of 49 and average follow-up time of 29.2 years), 11 studies assessed PA in late life (average age of 75.9 and average follow-up time of 5.9 years), and two assessed PA in adulthood without specification. For studies that assessed PA at mid-life, 2 out of 4 (50%) had statistically significant findings (p < 0.05), for studies that assessed PA during late life, 8 out of 11 (75%) had significant findings (p < 0.05), and 2 out of 2 (100%) of unspecified timing had significant findings (p < 0.05). Our review indicated that engaging in PA at least three times per week, for at least 15 minutes per session, was judged to be the minimum requirement tested for protective effects against AD in late life. Potential biological mechanisms were also discussed.

**Conclusion:** Our current review supports existing evidence that PA provides significant protection against the development of AD and found that the requirement of PA may be less than the current guidelines for sufficient and meaningful protection in late life. Excitingly, any form of PA tested can be protective against the development of AD, including household activities, suggesting that a wider variety of PA can be more appropriate for late life. More standardized and detailed studies need to update the benefits of PA, particularly in the areas of occupational, household/transportation, and age-group activities. Further research is needed to determine optimal PA thresholds in these groups.

## Introduction

Alzheimer’s Disease (AD), the most common type of neurodegenerative dementia, is a leading cause of mortality and morbidity among people with older age. It poses a significant burden on caregivers, families, and the health care system. As of 2024, an estimated 6.9 million Americans are living with AD, with 73% of them aged 75 and older and 33.4% aged 85 and older ^1^. If the current trends continue, the number of individuals living with AD could more than double to 13.8 million by 2060 ^1^. It has been estimated that about one-third or more of AD cases can be attributed to seven modifiable risk factors, including diabetes, midlife hypertension, midlife obesity, physical inactivity, depression, smoking, and education ^2,3^.

Physical activity (PA) likely contributes to the prevention or protection from the development of cognitive decline. Previous systematic reviews and meta-analysis data have shown that PA has a protective relationship against the development of AD ^4–6^. Additionally, PA has been linked to improving overall health and reducing the risk of cardiovascular disease, metabolic disease, and depression, suggesting that the protective benefit is likely multifactorial ^7–10^. Previous studies have also shown that PA improves cerebral perfusion, facilitates neurogenesis and synaptogenesis, and provides protective effects against β-amyloid accumulation and tau phosphorylation ^11,12^. These findings warrant further investigation into elucidating the specific association between PA and AD since PA is a cost-effective, time-effective, and often underutilized approach. Currently, there is no clear preventive strategy regarding the minimum amount of PA needed to provide a protective benefit against the development of AD. Therefore, this systematic review aims to elucidate these parameters to further clarify this protective association.

## Methods

### Literature Search

The systematic review followed PRISMA guidelines ^13^ and used Medline via PubMed and CINAHL databases. The search strategy involved medical subject headings and keywords like "exercise" OR "physical activity" AND "Alzheimer’s disease" OR "cognitive impairment" OR "dementia". We used a combination of these terms and filters to access the full search results. See supplemental figure 1 to see the detailed search strategy. The search was conducted in August 2023.

### Inclusion and Exclusion criteria

The inclusion criteria for the study are as follows: prospective cohort or interventional studies written in English that measured physical activity in a cohort of older adults without dementia, Alzheimer’s disease (AD), or cognitive decline at the beginning of the study, and measured the incidence of AD as an outcome. Reviews, case reports, cross-sectional studies, retrospective studies, and any other studies that did not meet the inclusion criteria were excluded.

### Study selection

Covidence software was used to screen the articles ^14^. Two reviewers, initials AS and AL, independently screened the articles by title and abstract, while a third reviewer with initials PL was consulted to address any discrepancies. Following this initial screening, the selected studies underwent a full-text review in accordance with the specified inclusion and exclusion criteria.

### Risk of Bias

The risk of bias was assessed using the Robins-E tool to assess the risk of bias in non-randomized follow-up studies of exposure effects ^15^. Studies were assessed according to Robins-E criteria by independent reviewers in seven domains: confounding, measurement of exposure, selection of participants, post-exposure intervention, missing data, measurement of the outcome, selection of reported result, and overall bias judgment. Figures were made using the Robvis tool^16^.

## Results

Our literature search yielded 2322 articles relevant to our research question. Following the initial title and abstract screening, 47 studies underwent a full-text review based on the inclusion and exclusion criteria. We excluded 26 studies for measuring the wrong outcome, 2 for the wrong study design, 1 for the wrong intervention, and 1 for not having the full text available. A total of 17 studies were selected for data retrieval, as shown in Figure 1. Two reviewers independently conducted data collection of study characteristics and outcome data of hazard ratios and odds ratios.

**Figure 1.**
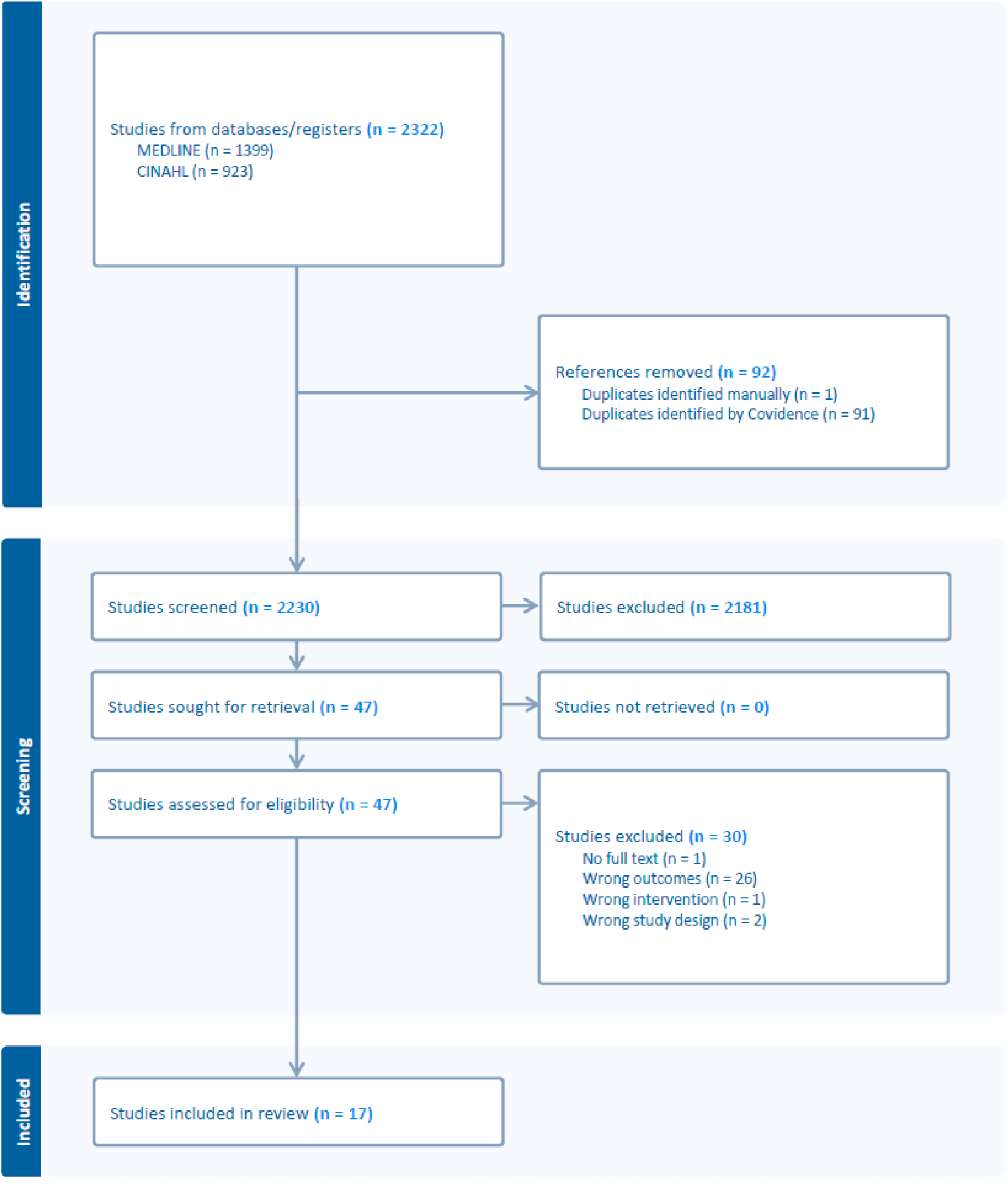
Flow Diagram depicting the study selection process

### Study Characteristics

Study characteristics were separated into those studying PA during mid-life, late life, and unspecified time of life (Tables 1, 2, 3). Overall, the study populations were from the USA, Canada, and various countries in Europe, including Sweden, Finland, France, Germany, Denmark, Italy, and the United Kingdom. 16 studied the general population, while one focused on patients. Sample sizes varied from 466 to 90,320 for a total of 206,463 participants. The average age was 67.7 years, and the follow-up times varied from 3.9 years to 44 years. Our method identified 92 duplications compared to 3,580 in a prior review ^6^, which seems to be due to the efficiency of machine-learning-assisted literature screening. 13 of the 17 studies reported a statistically significant inverse association between physical activity and the incidence of AD.

**Table 1:**
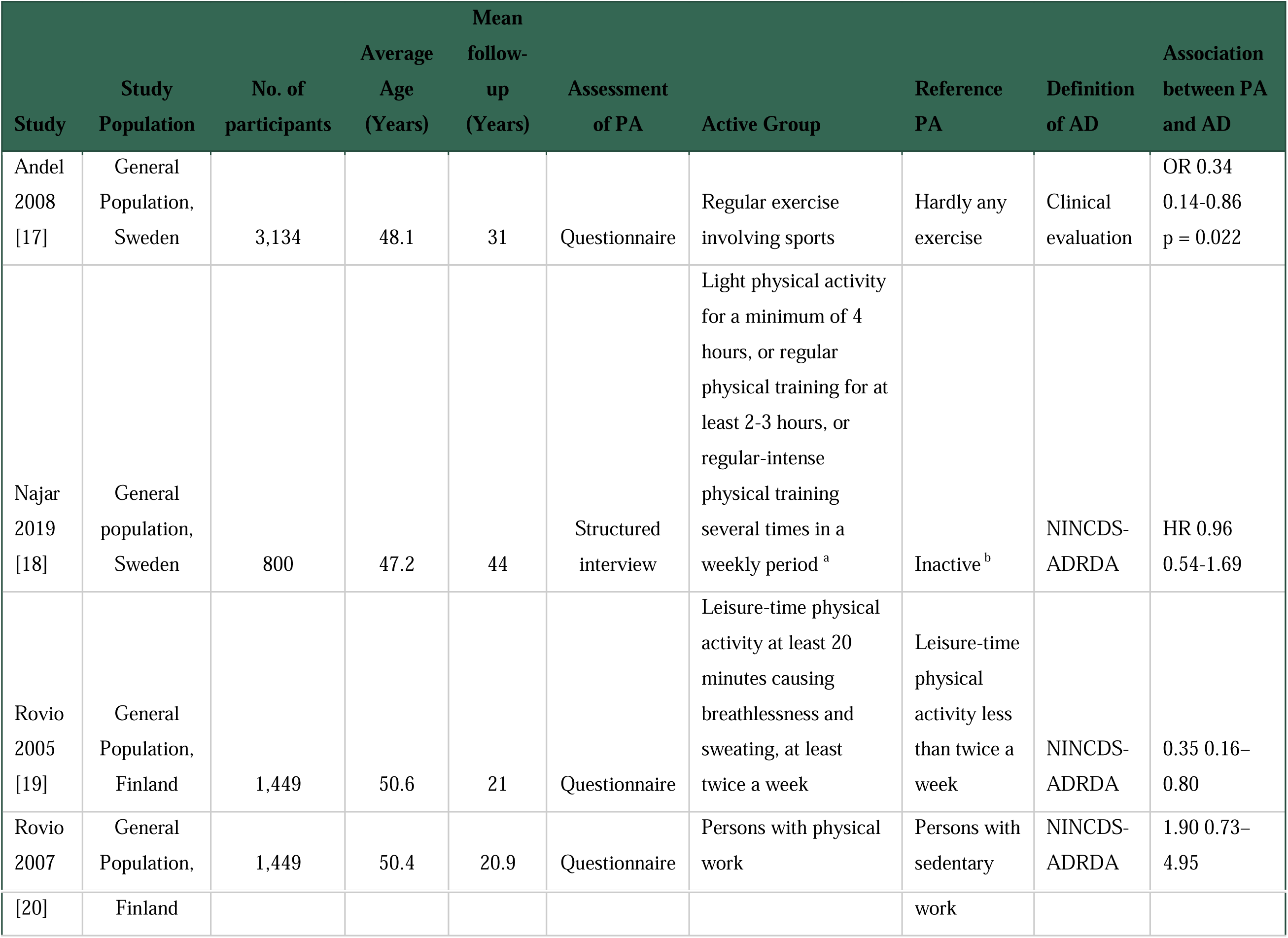
Study Characteristics, Mid-Life.

| Study | Study Population | No. of participants | Average Age (Years) | Mean follow-up (Years) | Assessment of PA | Active Group | Reference PA | Definition of AD | Association between PA and AD |
| --- | --- | --- | --- | --- | --- | --- | --- | --- | --- |
| Andel 2008 [17] | General Population, Sweden | 3,134 | 48.1 | 31 | Questionnaire | Regular exercise involving sports | Hardly any exercise | Clinical evaluation | OR 0.34<br>0.14-0.86<br>p = 0.022 |
| Najar 2019 [18] | General population, Sweden | 800 | 47.2 | 44 | Structured interview | Light physical activity for a minimum of 4 hours, or regular physical training for at least 2-3 hours, or regular-intense physical training several times in a weekly period <sup>a</sup> | Inactive <sup>b</sup> | NINCDS-ADRDA | HR 0.96<br>0.54-1.69 |
| Rovio 2005 [19] | General Population, Finland | 1,449 | 50.6 | 21 | Questionnaire | Leisure-time physical activity at least 20 minutes causing breathlessness and sweating, at least twice a week | Leisure-time physical activity less than twice a week | NINCDS-ADRDA | 0.35 0.16–0.80 |
| Rovio 2007 | General Population, | 1,449 | 50.4 | 20.9 | Questionnaire | Persons with physical work | Persons with sedentary | NINCDS-ADRDA | 1.90 0.73–4.95 |
| [20] | Finland |  |  |  |  |  | work |  |  |

**Table 2:**
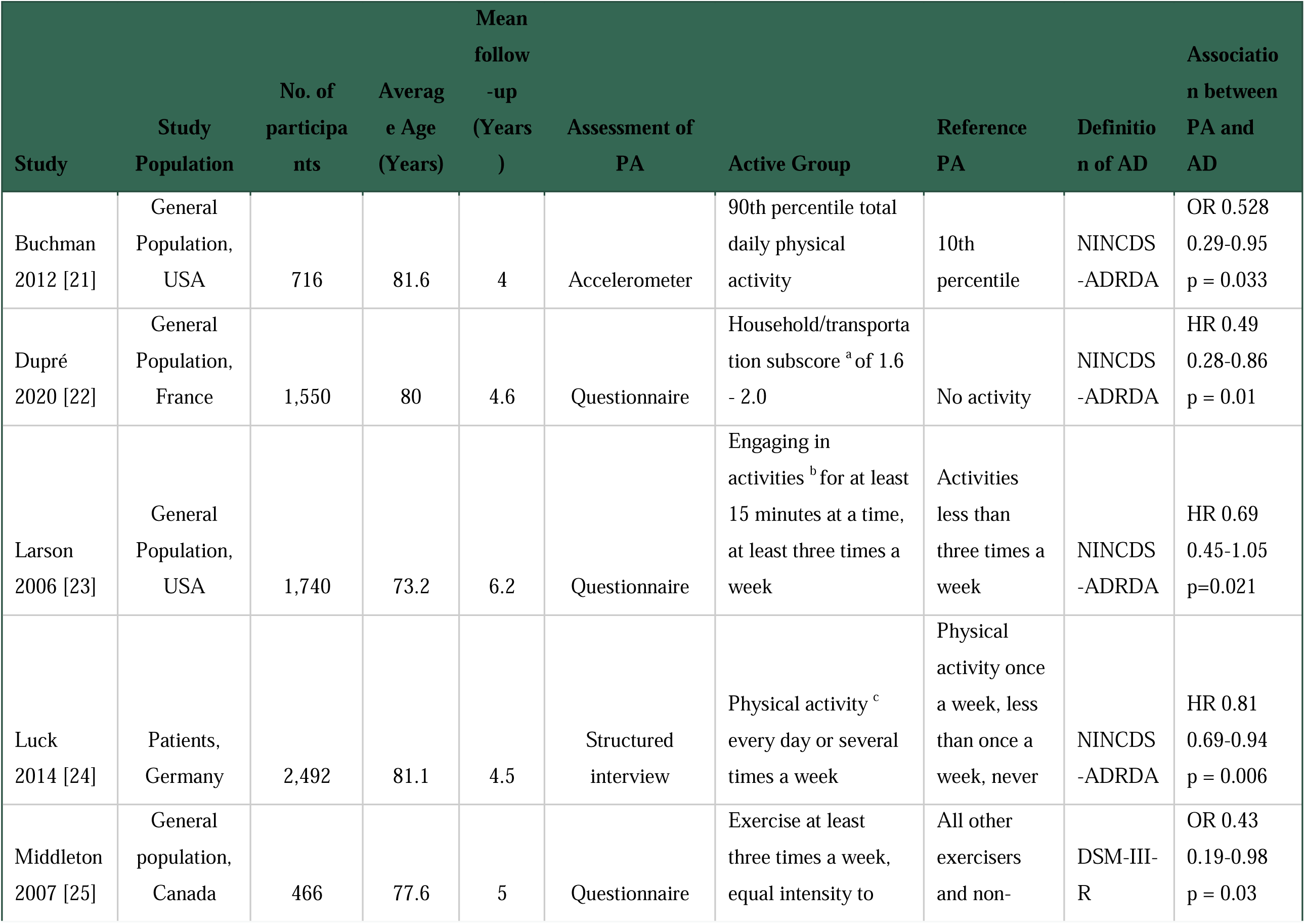
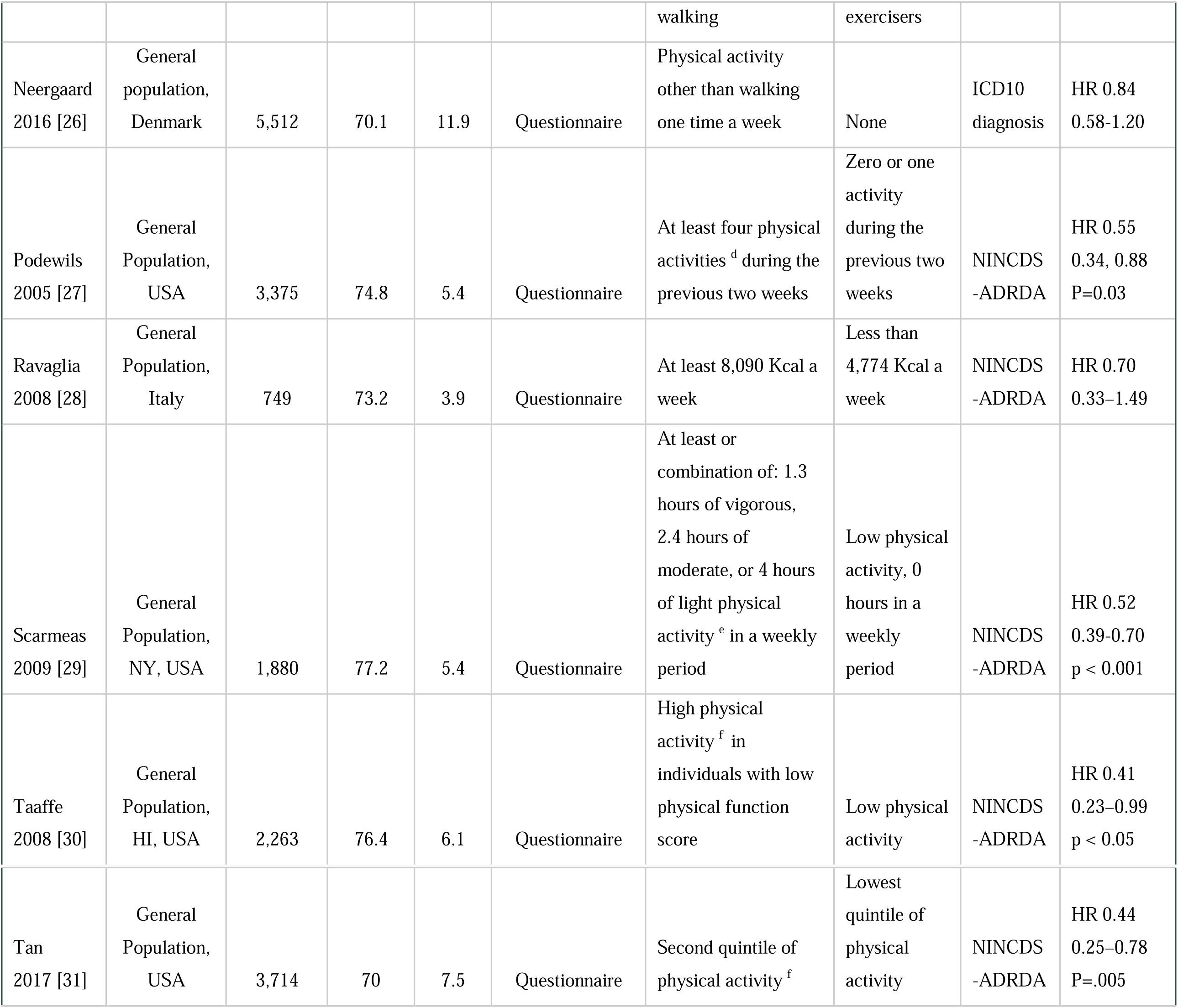
Study Characteristics, Late Life.

### Mid-life

Four studies measured PA during mid-life, as shown in Table 1.

The study populations were general populations of Sweden and Finland, with an average age of 49.1 years old and an average follow-up time of 29.2 years. Three studies measured PA with questionnaires ^17,19,20^, and one conducted structured interviews ^18^. Two out of four had significant findings that regular PA at midlife has a protective effect against AD ^17,19^. Two of the three studies measuring leisure-time PA were significant ^17,19^, and one measured occupational PA was not significant ^20^. Najar et al. (2019) and Rovio et al. (2005) included a measurement of intensity and duration in their thresholds of PA ^18,19^. Rovio et al. (2005) reported a significant decrease in the incidence of AD with physical activity with associated breathlessness and sweating for at least 20 minutes at a time and at least twice a week, compared to less than twice a week. Najar et al. (2019) did not report a significant relationship with a threshold of light PA of a minimum of 4 hours, regular physical training for at least 2-3 hours, or intense physical training several times per week.

### Late Life

#### 11 studies measured PA during late life, as shown in Table 2

The study participants comprised of general populations of USA, Canada, France, Italy, and Denmark, and a patient population in Germany. The study populations had average age of 75.9 years old and average follow-up time was 5.9 years. One study assessed PA with accelerometer data ^21^, one conduced structured interviews ^24^ and nine used questionnaires ^22,23,25–31^. Eight out of 11 (75%) studies reported significant findings ^21–25,27,29,31^. Six studies measured leisure PA ^23–27,29^ and four reported a significant relationship ^23–25,27,29^. One measured household/transportation PA and was significant ^22^. Four measured total PA ^21,28,30,31^ and three reported a significantly protective benefit ^21,30,31^.

Several studies included a parameter of duration or intensity in their PA assessments. Both Larson et al. (2006) and Middleton et al. (2007) found that PA at least three times a week conferred a significant protective benefit, but the former required least 15 minutes at a time, while the latter required an intensity equal to walking ^23,25^. Luck et al. (2014) found similar results with a threshold of PA every day or several times a week ^24^. Scarmeas et al. (2009) found that the combinations of light, moderate, or vigorous exercise several times a week decreased incidence of AD, compared to those who did low activity or 0 hours of physical activity ^29^.

Podewils et al. (2005) observed that at least four physical activities in a two-week period confers a significant benefit, compared to one or less physical activities ^27^.

Studies also calculated subscores and percentiles to compare levels of PA. Dupre et al. (2020) calculated a household or transportation subscore and found that the intermediate score conferred a protective benefit, compared to no activity at all ^22^. Two studies reported 24-hour physical activity calculated by average hours per day spent in different basal states. Taaffe et al. (2008) found that a high physical activity subscore was protective from developing AD, compared to a low physical activity score ^30^. Similarly, Tan et al. (2017) found that the second quintile of physical activity was protective, compared to the lowest quintile ^31^. Buchman et al. (2012) used accelerometer data for total daily physical activity and found the 90th percentile of PA had a protective benefit, compared to the 10th percentile of accelerometer data ^21^.

### Unspecified

Two studies analyzed a population in the United Kingdom that did not specify a targeted age range. The study populations had an average age of 60.1 years old (ranging from 37-73 years old) and an average follow-up time of 6.6 years (Table 3). Both studies used accelerometer data to assess total daily PA. Peterman-Rocha et al. (2021) and Zhong et al. (2023) used 900 metabolic equivalents per minute per week and 27 milli-gravity units per week, respectively, for the active group threshold ^32,33^. Both studies conferred a protective benefit at these thresholds.

**Table 3:** Study Characteristics, Unspecified time of life.

| Study | Study Population | No. of participants | Average Age (Years) | Mean follow-up (Years) | Assessment of PA | Active Group | Reference PA | Definition of AD | Association between PA and AD |
| --- | --- | --- | --- | --- | --- | --- | --- | --- | --- |
| Petermann-Rocha 2021 [32] | General population, UK | 84,854 | 64 | 6.3 | Accelerometer | Total physical activity intensity and duration of at least 900 MET/min/week | Less than 300 MET/min/week | ICD10 diagnosis | HR 0.26<br>0.12-0.55<br>P<0.001 |
| Zhong 2023 [33] | General population, UK | 90,320 | 56.2 | 6.9 | Accelerometer | Total physical activity average over 27 milli-gravity (milli-g) units/week | Total physical activity average over less than 27 milli-gravity (milli-g) units/week | ICD10 diagnosis | HR 0.74<br>0.60, 0.90<br>p = 0.003 |

### Risk of Bias Assessment

The risk of bias assessment in the overall quality of evidence was “low risk” in 13 studies and “some concerns” in four studies (Figure 2). The studies were placed in the “some concerns” category due to potential bias in confounding, measurement of exposure, and missing data. None of the studies were deemed high risk or very high risk of bias.

**Figure 2:**
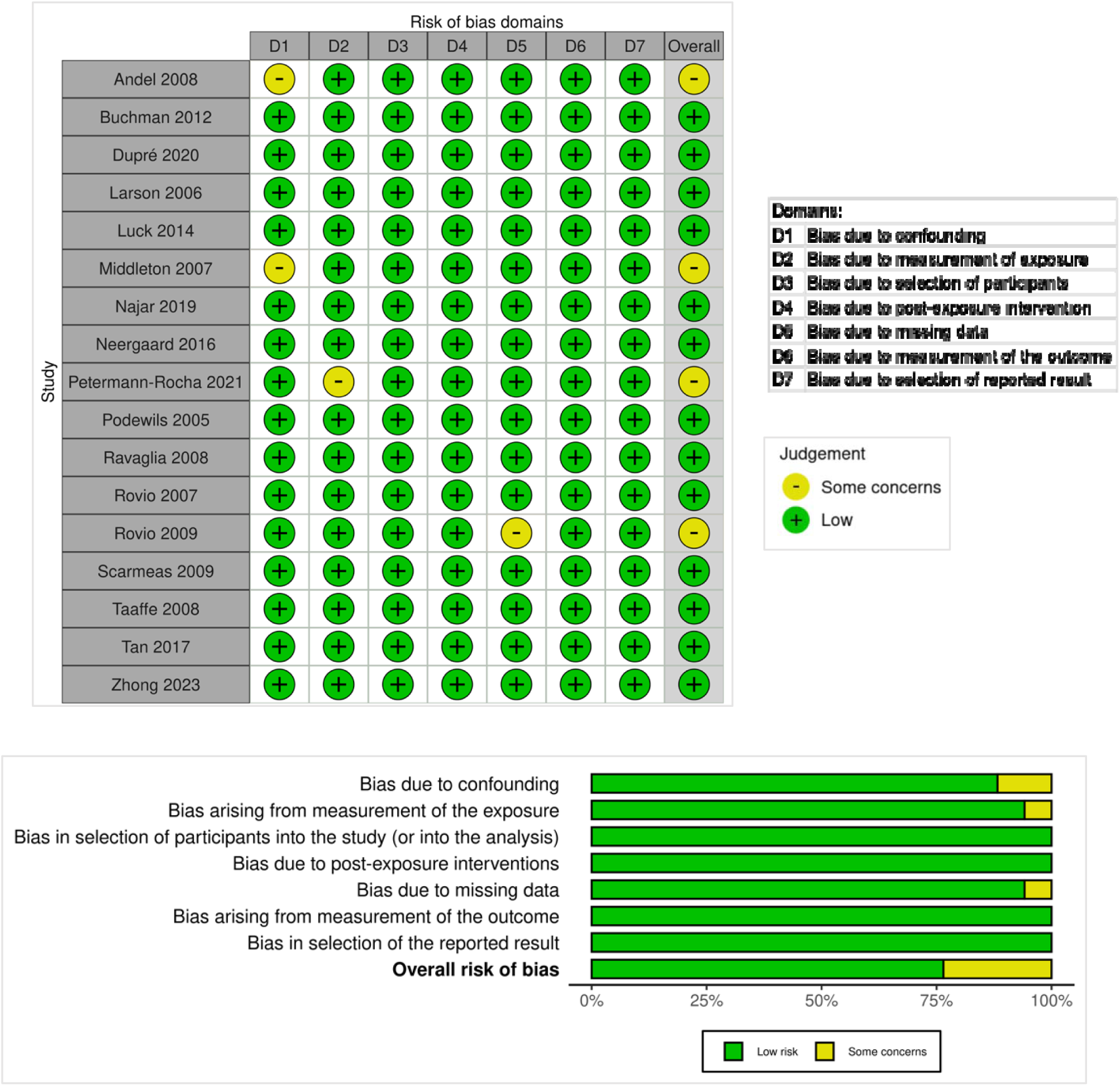
Risk of Bias.

## Discussion

Our current review incorporated a wide range of studies with a low risk of bias that examined the relationship between PA and AD in different life stages. While we found evidence that PA protects against developing AD in both the mid-life and late lifetime period, we found that the protective effect of PA in late life has been consistently significant. We will now discuss these findings in turn, as well as potential biological mechanisms.

### Mid-life

Two out of the four included studies that assessed mid-life PA resulted in a significant protective benefit from developing AD. Three studies— Andel et al. (2008), Najar et al. (2019), and Rovio et al. (2005) —measured leisure time PA at mid-life, with at least two decades of follow-up. ^17–19^. Of these, only Najar et al. (2019) did not find significant protection from AD, although the study did report a reduced risk of mixed dementia and dementia with cerebral vascular disease ^18^.

In contrast, both Andel et al. (2008) and Rovio et al. (2005) presented a strong significant reduction in AD risk ^17–18^. Andel et al. (2008) specifically assessed regular exercise involving sports from age 25 to 50 years old ^17^, which highlights the significant benefit of cumulative PA leading up to mid-life. This concurs with a previous meta-analysis data which concluded that PA provides a significant risk reduction, especially when comparing moderate levels of PA to extreme sedentary behavior ^4,6,34^. However, Najar et al. (2019) did not find a significant difference between light PA (such as walking) and inactivity ^18^, suggesting that this level of activity may not be sufficient to offer significant protective benefits. In contrast, Rovio et al. (2005) assessed leisure-time PA with more specific criteria: at least 20 minutes of activity that caused breathlessness and sweating, performed at least twice a week ^19^. This study’s inclusion of a measure of intensity in their definition of PA may provide valuable insights into the minimum intensity required for a protective effect, with breathlessness and sweating potentially serving as indicators of more protective activity levels.

One study measured mid-life occupational PA and determined it did not provide a significant protective effect against AD ^20^. This suggests that leisure-time activity and occupational activity have potentially different effects on the development of AD. There are many hypotheses about why this is the case, with some evidence that the population who worked occupations requiring manual labor are associated with lower socioeconomic status and more adverse health outcomes after retirement, including dementia and cardiovascular disease ^35–37^. However, in the study, Rovio et al. (2007) found that occupational PA did not demonstrate a statistically significant result, even when controlled for socioeconomic variables, including income and education levels ^20^. Leisure time PA was also controlled for, which suggests that occupational PA likely does not have an independently significant protective effect on the incidence of AD. More studies are needed regarding these nuances.

### Late life

The current analysis showed that physical activity, especially during later life, resulted in a significant protective effect against developing AD. This supports previous studies indicating that PA has a protective effect on the development of neurodegenerative disease ^38,39^, AD specifically ^3,4,6,37,40–42^, and among older adults ^42,43^. Our review found that various types of PA are protective, including household or transportation, leisure, sports, or even total daily activity measured by accelerometer.

Thus far, there has been a lack of a clear consensus on specific practical recommendations for PA ^6^, and our review provides clearer guideline recommendations for late life intervention.

While PA methods and thresholds vary, we conclude that engaging in PA at least three times per week, for at least 15 minutes per session, can be a suggestion for the minimum threshold for protection against AD in late life. Studies by both Larson et al. (2006) and Luck et al. (2014) indicate that engaging in PA at least three times a week, compared to less frequently, offers a significant protective effect ^23,24^. These findings are in congruence with Podewils et al. (2005), who reported significant protection for individuals engaging in at least four physical activities in a two-week period ^27^. Although Neergard et al. (2016) found that PA at least once a week did not confer a significant prevention from AD, the study did find that PA at least three times a week did confer a significant benefit from preventing all-cause dementia and vascular dementia ^26^.

Taken together, these studies suggest that a frequency of at least three times a week of PA is likely necessary for meaningful protection against the development of AD.

The most recent Physical Activity Guidelines for Americans by the US Department of Health and Human Services recommend a combination of frequency and intensity of PA per week. The recommendation for adults entails at least 150 minutes to 300 minutes a week of moderate-intensity (such as brisk walking), or 75 minutes to 150 minutes a week of vigorous-intensity aerobic physical activity (such as jogging or running), or an equivalent combination of moderate or vigorous activity, as well as muscle strengthening exercises at twice two times a week ^44^. The guidelines are rooted in decades of substantial research aimed to provide broad protection from a large range of health risks- including cardiovascular diseases, several types of cancers, dementia, anxiety and depression, and all-cause mortality ^44,45^.

Unfortunately, only about a quarter of Americans reach these recommended levels of activity ^45^. However, there is evidence that even a frequency of three times a week, with even mild to moderate intensity, can provide substantial benefits. A recent study identified a minimal amount of 40 minutes per week of vigorous PA can reduce Alzheimer’s disease-related mortality, with an optimal duration of 140 minutes a week ^46^. Remarkably, the researchers found that even 40 minutes a week can prevent 12,238 deaths from Alzheimer’s per year ^46^. These findings align with the broader consensus that while 140-150 minutes of higher-intensity PA is optimal, many older adults can have significant benefits from a smaller amount.

Further, if public health authorities or clinicians can recommend for adequate physical activity as a form of prevention that can be beneficial even over the age of 65, it can encourage those in late life to continue to exercise and adopt “never too late” mentality on preventing disease ^42^. Three of the included studies had an average age of over 80 years old and still had a significant benefit ^21,22,24^, with a variety of activities, even household chores ^22^. This suggests that routine activities that are inclusive of the capabilities in the older population can have significant benefits.

Additionally, given that there is less clarity regarding the relationship between physical activity and improving cognitive outcomes after developing cognitive decline or dementia ^47,48^, the emphasis should shift to using physical activity as a key preventative therapy. This recommendation can benefit patients at any time, as physical activity is relatively accessible, safe, and low-cost.

### Potential Biological Mechanisms

The protective benefit of exercise in preventing the development of Alzheimer’s is undoubtedly multifaceted. It is estimated that about 35% of dementia cases are attributable to nine risk factors encompassing elements of socioeconomic status, chronic diseases such as hypertension, diabetes, obesity, hearing loss, depression, diabetes, and modifiable risk factors such as smoking and physical inactivity ^49^. PA was found to elevate various neurochemicals, including growth hormone (GH), insulin-like growth factor-1 (IGF-1), and brain-derived neurotrophic factor (BDNF), which play key roles in multiple biological processes ^50^. Namely, increased basal IGF-1 was found to have an inverse correlation with neurocognitive decline measured by the mini-mental state examination (MMSE) ^51^. IGF-1 is a hormone linked to neurogenesis in the hippocampus, and notably, alterations in its levels often precede the development of diabetes ^52^. PA has been demonstrated to be beneficial for these chronic conditions, suggesting a shared network of beneficial effects ^37,53^.

Additionally, there is evidence that there is a complex interplay between lifestyle interventions and the genetics of age-related diseases. Previous studies have discussed the genetic pathways leading to common age-related chronic illnesses, including diabetes, hypertension, and other cardiovascular diseases that are genetically related to the development of AD ^54,55,56^. Given physical activity has long been known to be protective against the development of all these illnesses, its benefits across several domains of health are likely intertwined and compounded ^37^. Specifically regarding APOE 4 and 2 alleles, which have long been known as strong genetic markers for AD, there is some evidence that lifestyle and environmental factors can affect the risk of expression ^27,57,58^. APOE 4 dose was found to modify the effect of pulse pressure on cerebral blood flow (CBF) in memory-related brain regions, including the hippocampus, entorhinal cortex, and inferior parietal cortex ^59^. Higher CBF in the entorhinal cortex was associated with greater memory stability in those who were 4 carriers ^60^. Given cerebral hypoperfusion is an early and persistent feature in AD progression ^61,62^, these findings suggest that targeted exercise interventions may help improve cerebral blood flow and mitigate vascular contributions to cognitive decline. Similarly, greater regional CBF decline was found in 4 carriers when compared to non-carriers ^63^. Additionally, APOE levels in certain genotypes have been shown to be inversely related to PA in preliminary studies in young adults, and young carriers of APOE4 can benefit from physical exercise and lifestyle modification ^58,64^. More research is needed in this area to further elucidate the relationship between PA and AD on a genetic level.

### Strengths and Limitations

Strengths of the current analysis include the inclusion of studies with large cohorts with sufficient years of follow-up and the completion of bias assessment that ensured the quality of included studies. Our review was the first to provide practical guidelines for optimal PA levels that can be protective of developing AD in late life.

This present study is not without limitations. The studies included had sizable heterogeneity. Each study employed unique parameters for statistical analysis and different reference values, making head-to-head comparisons and collective analysis difficult. For example, while calculated sub-scores, percentiles, and quintiles are useful for independent analysis, it can be difficult to create guidelines regarding minimum requirements from this type of data.

Assessment of PA levels also varied, with some studies using accelerometer data, and some using self-report questionnaires or interviews. Self-report data is susceptible to self-report bias and social desirability bias. While free from self-report bias, the accelerometer could be susceptible to the Hawthorne effect while the patients were monitored. However, overall, the biases of these studies were assessed and none were high risk. Another limitation includes the lack of a longitudinal measure of PA. Most of the studies included in this present review assessed PA at a cross-sectional time point, making it difficult to assess if subjects were experiencing benefits from PA over a short time period or a cumulative effect over time. It has been stated that the benefit of PA was only associated with the protection of a four-year follow-up period ^64^. Therefore, studies can benefit from more years of follow-up, with levels of PA assessed at more frequent time periods, to provide further insight into the protective benefits.

## Conclusion

In conclusion, this systematic review highlights the significant protective effect of PA against the development of AD, particularly leisure-time PA during late life. Our findings underscore that late life PA performed at least three times per week for a minimum of 15 minutes per session, can significantly reduce the risk of developing AD. Encouraging older adults to adopt a routine that includes regular physical activity, even in modest amounts, could be an important public health strategy to mitigate the growing burden of AD in aging populations. Additionally, it is important for future studies to focus on longitudinal and less heterogeneous assessments of PA to provide clearer guidelines for the minimum threshold of PA needed for protective benefits.

## Data Availability

All data produced in the present work are contained in the manuscript

## Financial Disclosures

None reported

## Support

None reported

## Ethical Approval

N/A

## Informed consent

N/A

## Acknowledgment

We thank the members of Murakami laboratories for helpful discussion and technical assistance. The manuscript has been revised for grammar by the members. It was also revised using Grammarly embedded with AI assistance: https://www.grammarly.com (Last accessed on March 31, 2025) for a minor grammar and spelling check.

## Appendix: Key Word Search Strategy for MEDLINE and CINHAL

Pubmed via MEDLINE

("Alzheimer Disease"Mesh OR “Alzheimer’s Disease”Text word OR “Alzheimer Disease”Text word OR "Cognitive Dysfunction"Mesh OR “Cognitive Impairment”Text word OR “Cognitive Dysfunction”Text word OR "Dementia"Mesh OR "Dementia"Text word) AND ("Exercise"Mesh OR "Exercise"Text word OR "Physical Activity"Text word)

*Filters Applied: Clinical Study, Clinical Trial, Comparative Study, Controlled Clinical Trial, Observational Study, Randomized Controlled Trial*

CINHAL

(MM "Alzheimer’s Disease" OR MM "Delirium, Dementia, Amnestic, Cognitive Disorders" OR TI "Alzheimer’s" OR TI "Alzheimer" OR TI "Dementia" OR TI “Cognitive Dysfunction” OR TI “Cognitive Impairment”) AND (MM "Exercise" OR MM "Physical Activity" OR TI "Exercise" OR TI "Physical Activity

## Notes

### Competing Interest Statement

The authors have declared no competing interest.

### Funding Statement

This study did not receive any funding

### Author Declarations

The study used (or will use) ONLY openly available human data that were originally located at: MEDLINE: https://www.medline.com/ CINAHL: https://www.ebsco.com/products/research-databases/cinahl-database

